# Population genomic screening leads to improved lipid management in patients with familial hypercholesterolemia

**DOI:** 10.1101/2025.03.13.25323900

**Authors:** Matthew E. Levy, Kelly M. Schiabor Barrett, Megan N. Betts, David Kann, Alexandre Bolze, Basil Khuder, Natalie Telis, Lisa M. McEwen, Chad Haldeman-Englert, Jeremy Cauwels, Douglas Stoller, C. Anwar A. Chahal, Christopher N. Chapman, Ashley A. Waring, Douglas A. Olson, Joseph J. Grzymski, Nicole L. Washington, William Lee, Elizabeth T. Cirulli, Catherine Hajek

**Author notes:** Corresponding author: Matthew Levy, PhD. Helix. Mail: 400 S El Camino Real, Suite 300, San Mateo, CA 94402.

## Abstract

**Background:** The Helix Research Network^TM^ program is a large population genomics initiative that screens an all-comers population of patients for CDC Tier 1 genetic conditions, including familial hypercholesterolemia (FH). We evaluated changes in clinical management and LDL cholesterol (LDL-C) levels among patients we identified to have FH.

**Methods:** Participants across eight U.S. health systems provided samples that underwent clinical-grade exome sequencing. Individuals with a positive screening result for a Tier 1 condition were offered no-cost genetic counseling through their health system. Using medication and laboratory testing records, we evaluated changes in patients’ lipid-lowering therapies and LDL-C levels.

**Results:** Among 211,263 adults enrolled between 2017-2024, 1,020 (∼1/207) had a pathogenic FH variant in *LDLR* (72%), *APOB* (27%), or *PCSK9* (1%). Of the 453 with retrospective and prospective electronic health record (EHR) data available (mean of 13.6 and 2.3 years, respectively), 85% lacked a prior clinical FH diagnosis. Overall, 33% received new/modified lipid-lowering therapy within the first year, but this proportion was higher in those with a newly documented FH diagnosis code (55% vs 18% for those without documentation, p<0.001). Patients with new/modified therapies had a mean LDL-C reduction of 59 mg/dL, compared to 18 mg/dL in patients with no therapeutic change (difference=41 mg/dL, p<0.001).

**Conclusions:** Identification of patients with FH likely led to improvements in clinical management and LDL-C levels. EHR documentation of the diagnosis code was associated with greater likelihood of therapeutic modifications which, in turn, were associated with larger LDL-C reductions. Findings underscore the powerful potential of population genomic screening for promoting optimal lipid management in individuals with FH.

## Introduction

Familial hypercholesterolemia (FH) is a genetic disorder characterized by lifelong high serum levels of low-density lipoprotein cholesterol (LDL-C), which, if left untreated, contributes to an increased risk of early-onset atherosclerosis and cardiovascular events.^1^ Untreated males have a 50% risk of experiencing a coronary event by 50 years of age, and untreated females have a 30% risk of experiencing an event by 60 years of age.^2^ FH is typically inherited in an autosomal dominant pattern, involving pathogenic variants in genes central to lipid metabolism, including *LDLR* (LDL receptor), *APOB* (apolipoprotein B), and *PCSK9* (proprotein convertase subtilisin/kexin type 9).^3^ Additionally, a rarer autosomal recessive form is caused by homozygous mutations in *LDLRAP1* (LDL receptor adaptor protein-1).

Despite a global prevalence of approximately 1 in every 250 individuals,^4^ FH is underdiagnosed and undertreated. In the absence of genetic testing, FH is frequently overlooked in clinical practice, as hypercholesterolemia is common in the general population, making it challenging to distinguish FH from other causes of elevated cholesterol.^5^ Reflecting the high-risk nature of FH and the benefits of intensive therapy, treatment goals for FH are evolving toward more aggressive LDL-C targets, which would typically not be sought in the absence of an FH diagnosis. While some guidelines recommend LDL-C <100 mg/dL for primary prevention, there is a growing consensus to aim for <70 mg/dL in individuals with FH (or ≥50% reduction), especially for those with additional risk factors or established cardiovascular disease.^6,7^

With the growing integration of genomics into routine healthcare,^8^ large-scale screening programs are able to identify individuals with genetic variants associated with FH.^9–12^ Within the Healthy Nevada Project, 102 out of 26,906 participants (∼1 in 264) were found to have an FH-associated pathogenic variant in *LDLR*, *APOB*, or *PCSK9*, and only 14% of those would have been eligible for genetic testing based on guidelines.^10^ In the Geisinger MyCode project, none of the 93 patients with an FH-associated variant detected had previously received a clinical diagnosis.^9^ Based on the Simon Broome diagnostic criteria, a positive genetic test alone is sufficient for a definitive FH diagnosis,^13^ while, in slight contrast, the Dutch Lipid Clinic Network requires one additional form of clinical evidence (e.g., high LDL-C).^14^

Prior studies of population genomic screening programs have reported on the prevalence and risk associated with FH, but little is known about the impact of such programs on changes in clinical care and outcomes. The Helix Research Network^TM^ program^15^ is a large multi-center population genomics initiative that screens an all-comer population of adult patients for CDC Tier 1 genetic conditions, including FH.^16^ In this article, we evaluate the impact of the population screening program on the clinical management and LDL-C levels among patients identified to have an FH-associated pathogenic variant. Our impetus is to provide insights into the public health importance of genomic screening and its potential to enhance the care of patients with FH.

## Methods

### Participants

As part of the Helix Research Network, adult patients across eight health systems participated in a population genomics screening program.^15^ There were no inclusion or exclusion criteria pertaining to FH or other diagnoses related to FH. Participants provided saliva or blood samples that underwent Exome+^Ⓡ^ sequencing at Helix for subsequent evaluation of pathogenic/likely pathogenic (P/LP) variants for CDC Tier 1 genetic conditions, as previously described.^10,17^ Individuals with a positive screening result for one of the conditions, including FH, were offered no-cost genetic counseling through their health system. The health system’s study team initiated outreach to facilitate results disclosure, and genetic counselors discussed the implications of the positive test result with patients, including potential impacts on family members and options for family variant testing. Results were integrated into each participant’s electronic health record (EHR). In cases where initial contact attempts were unsuccessful, a certified letter containing a clinical action plan was sent to the participant. Following result disclosure and referral to genetic counseling, subsequent care was expected to follow established health system clinical workflows.

For research purposes, longitudinal EHR data were retrospectively collected for participating patients and transformed into the Observational Medical Outcomes Partnership (OMOP) Common Data Model (CDM) version 5.4.^18^ The participating protocols are DNA Answers (St. Luke’s University Health Network, Pennsylvania), GeneConnect (Cone Health, North Carolina), the Genetic Insights Project (Nebraska Medicine), the Healthy Nevada Project (Renown Health, Nevada), ImagineYou (Sanford Health, Upper Midwest), In Our DNA SC (Medical University of South Carolina), myGenetics (HealthPartners, Minnesota), and The Gene Health Project (WellSpan Health, Pennsylvania). Study protocols were reviewed and approved by their respective Institutional Review Boards (projects 956068-12 and 21143). All participants provided written informed consent prior to participation, and all data used for research were deidentified.

### Variant interpretation

Variant interpretation for *APOB* and *PCSK9* were limited to specific, well-established pathogenic variants for FH (Supplemental Data File 1). Gene-level variant interpretation for *LDLR* was completed for the entire Helix Research Network cohort (N=211,263) using a two-step approach. First, a variant was considered P/LP if it had a known and well-established clinical pathogenic interpretation (i.e., no variant of uncertain significance [VUS] or benign interpretations present in ClinVar across high volume laboratories, using search strings [’ClinGen’, ’Quest’, ’Sema4’, ’Natera’, ’Invitae’, ’All of Us’, ’Baylor’, ’GeneDx’, ’Ambry’, ’LapCorp’, ’Color’, ’Myriad’, ’Brigham’] and/or a P/LP interpretation by the Familial Hypercholesterolemia Variant Curation Expert Panel [FH VCEP]). For all remaining variants, the American College of Medical Genetics and Genomics and the Association for Molecular Pathology (ACMG-AMP) variant interpretations were completed programmatically following the gene-specific scoring recommendations from the FH VCEP. Data from case studies as well as patient-specific information such as presenting symptoms or family history were not considered for these interpretations, given the population screening context as opposed to diagnostic context. All variants seen in the Helix Research Network, relevant annotations, scoring by ACMG-AMP data category, and resulting interpretation based off of point totals (pathogenic [>5], higher scoring VUS [3 to 5], lower scoring VUS [-1 to 2], benign [< -1]) are available in Supplemental Data File 1. Variant annotations were made based on the Matched Annotation from NCBI and EMBL-EBI (MANE) transcript for each gene and leveraged the following tools: VEP-104, GnomADv4, REVEL, SpliceAI, and the ClinVar database (accessed: 11/20/2024); and case-control (ACMG-AMP criterion PS4) data were obtained from systematic variant-level association tests internally-calculated using LDL-C levels from the UK Biobank and All of Us cohorts, with statin use included as a covariate. Variants were re-interpreted in the deidentified research dataset, with P/LP assertions confirmed by a clinical laboratory.

### Analyses

For this analysis, we evaluated multiple EHR-based clinical outcomes within up to three years post-screening among patients identified to have an FH-associated P/LP variant. We limited the analysis to those with ≥6 months of both retrospective and prospective data pre- and post-genetic screening, respectively. The date that biospecimens were received by Helix’s laboratory was used as the screening date. First, we evaluated whether patients received a documented diagnosis code for FH (SNOMED 398036000 or ICD-10-CM E78.01). Second, we assessed whether patients received new or modified prescriptions for LDL-lowering therapies including statins, ezetimibe, PCSK9 inhibitors, bile acid sequestrant, inclisiran, and bempedoic acid. Statins were classified as low-, moderate-, or high-intensity based on the statin type and dosage according to American Heart Association guidelines.^6^ Third, we computed changes in serum LDL-C concentrations, comparing each follow-up LDL-C result to the most recent LDL-C result pre-screening (within up to 1 year earlier) or, alternatively, to the earliest LDL-C result through up to 60 days post-screening (considered the baseline value, as available, in instances with no result in the 60 days before screening). Continuous differences as well as proportions of patients achieving targets of <70 mg/dL and ≥50% reductions were calculated. The OMOP CDM concept sets used to define LDL-lowering agents and EHR-based diagnosis measures are available in Supplemental Data File 2.

Patients’ demographic and clinical characteristics were compared between participants identified to have an FH-associated P/LP variant and those in the broader Helix Research Network. The proportions with new or modified LDL-lowering therapies post-screening were compared between patients with and without documentation of a clinical FH diagnosis code using chi-square tests. Rates of treatment with new or modified LDL-lowering agents by month since screening were also compared between patients with positive vs negative FH screening results, with monthly proportions calculated among patients with available follow-up for the full month’s duration (and with no new or modified therapy having already occurred in an earlier month). Additionally, proportions achieving LDL-C targets and mean maximum changes in LDL-C were compared between patients with and without therapeutic changes using chi-square testing and t-tests. Analyses were performed using R software, version 4.2.3.

## Results

### Prevalence of FH-associated P/LP genetic variants

Within the Helix Research Network, a cohort of 211,263 adults underwent screening for FH and other CDC Tier 1 genetic conditions between January 2017 and November 2024, with 48.6% of them having occurred during 2024. Overall, 1,020 individuals (0.48% or approximately 1 in 207) were found to carry a P/LP variant linked to FH. The majority of those patients with FH had a P/LP variant in *LDLR* (731 of 1,020, or 71.7%), while 277 (27.2%) had a P/LP variant in *APOB* (with the variant p.Arg3527Gln accounting for 89.9% of these) and 12 (1.2%) had a P/LP variant in *PCSK9*. All patients with FH were heterozygotes, with the sole exception of one person with a homozygous *LDLR* variant. There were no additional detections of homozygous FH or compound heterozygous FH, and there were no patients with a homozygous or compound heterozygous *LDLRAP1* variant.

### Participants with P/LP variants had high LDL-C prior to FH screening

Out of the 1,020 patients with FH, EHR data were available for 777 (76.2%). Among these, 453 patients (58.3%) had medical visit(s) both ≥6 months before and after their genetic screening (as of the dates of data extraction), and thus qualified for inclusion in this analysis.

Similar to the broader network, the mean age of FH patients was 52 years, and 72.4% were female (Table 1). The mean body mass index was 29.3 kg/m^2^, 37.5% had a hypertension diagnosis, and 13.0% had a type 2 diabetes diagnosis. Compared to participants without an FH-associated P/LP variant, those with a P/LP variant had higher mean serum concentrations of LDL-C based on both the most recent pre-screening measurement (143 mg/dL vs 105 mg/dL) and highest historical measurement (193 mg/dL vs 127 mg/dL). Patients with FH were more likely than those without FH to have been previously treated with an LDL-lowering agent (61.6% vs 27.5%) and to have a prior coronary artery disease (CAD) diagnosis (14.8% vs 6.3%). Only 21.9% of patients with FH had LDL-C <100 mg/dL prior to screening, even though 75.6% of them had received LDL-lowering therapy in the prior year, and fewer (6.7%) had LDL-C <70 mg/dL (not shown).

**Table 1.**
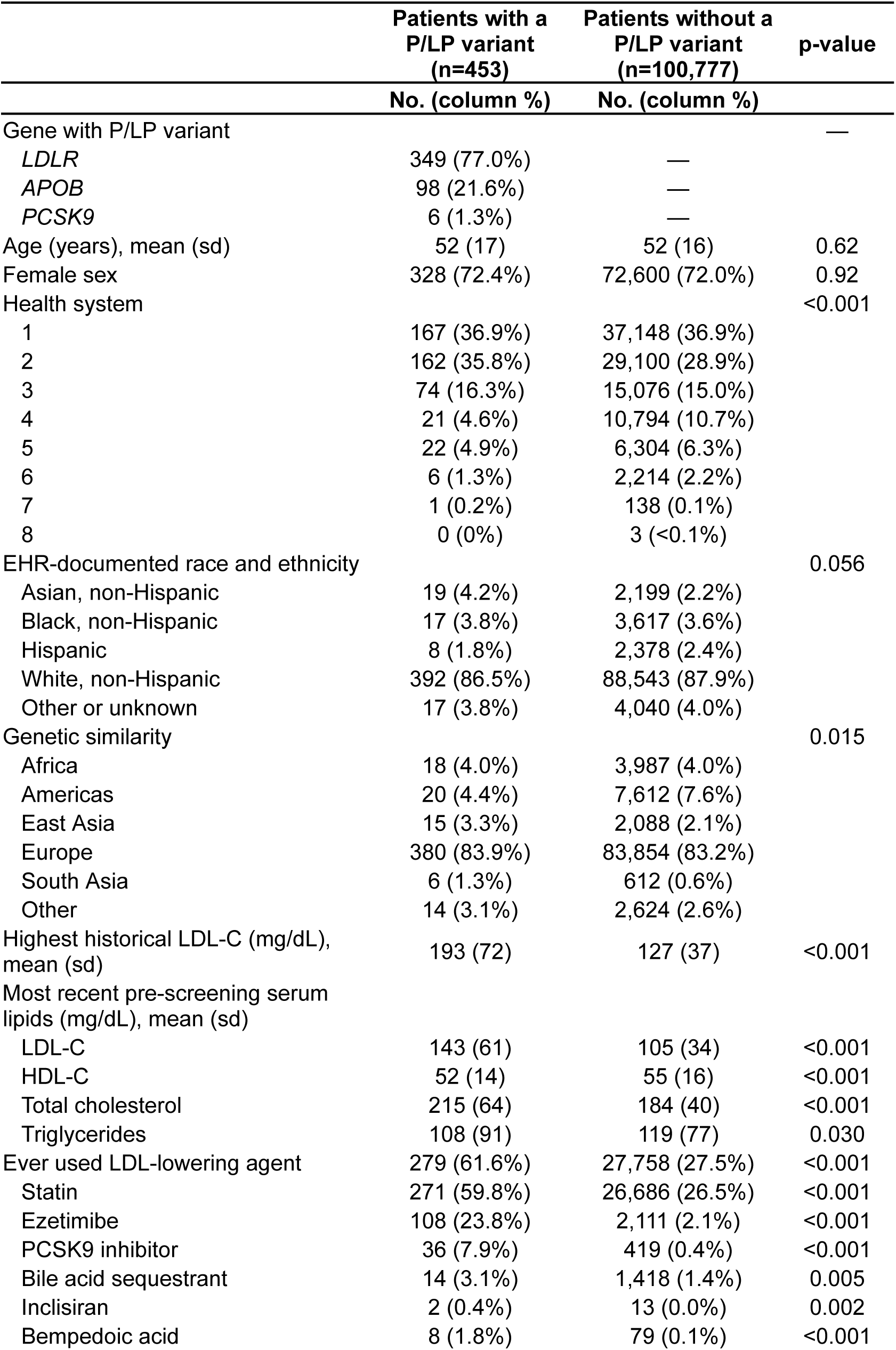
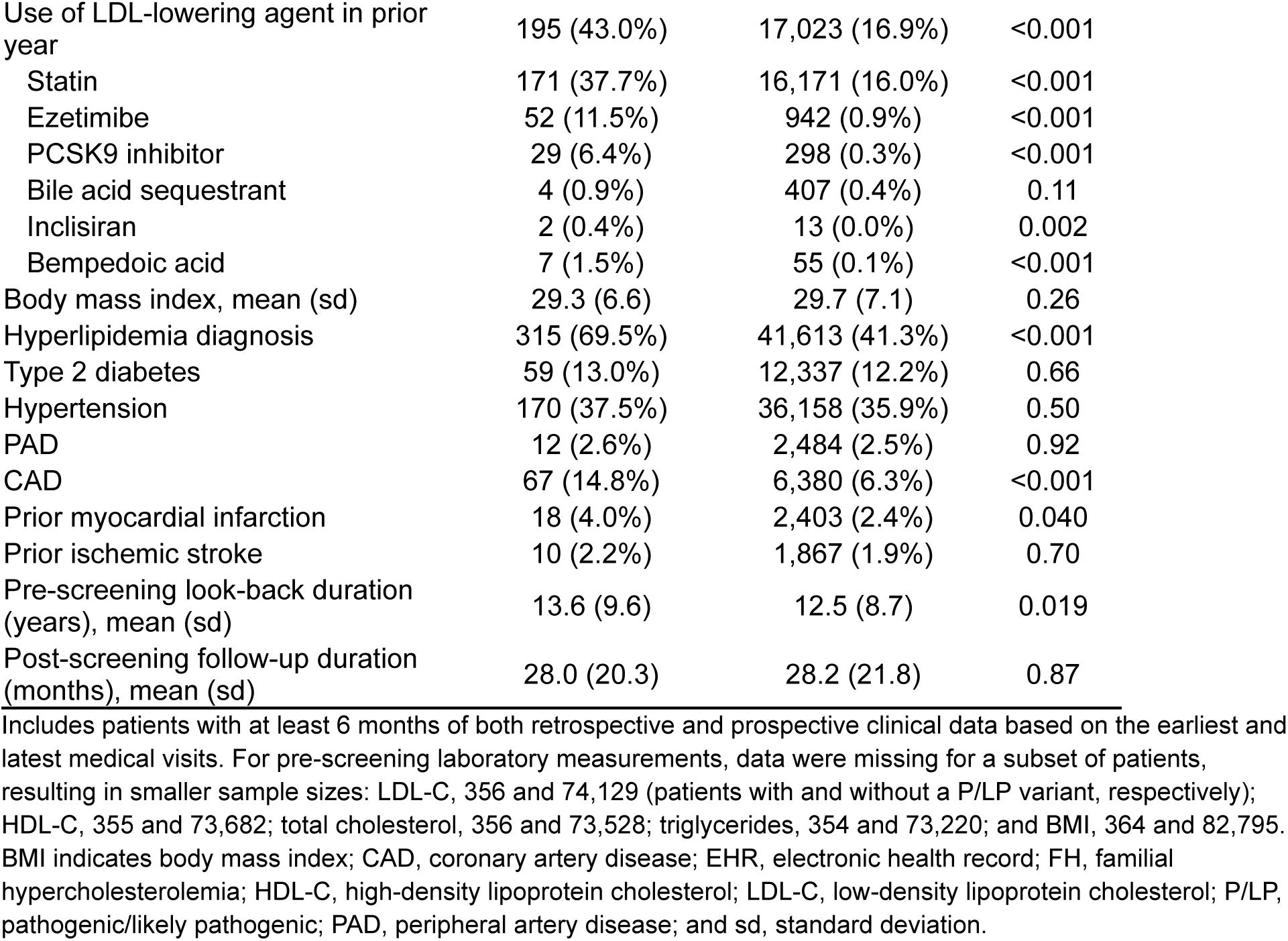
Characteristics of Helix Research Network participants with and without an FH-associated P/LP variant.

### Prior documentation of a clinical FH diagnosis and LDL-lowering therapy

The majority of patients with an FH-associated P/LP variant (84.8%) did not have a prior clinical diagnosis of FH documented in the EHR, although 69.5% had been diagnosed with general hyperlipidemia. Those with a prior clinical FH diagnosis were more likely than those without the diagnosis to have used an LDL-lowering agent within the previous year (66.7% vs 38.8%, p<0.001; Figure 1A). When stratifying by baseline CAD diagnosis status, this pattern was observed among patients without CAD (64.7% vs 34.3%, p<0.001), but not among those with CAD, who were largely treated irrespective of prior FH clinical diagnosis status (72.2% vs 69.4%, p=1.0) (Figures S1A and S1B).

**Figure 1.**
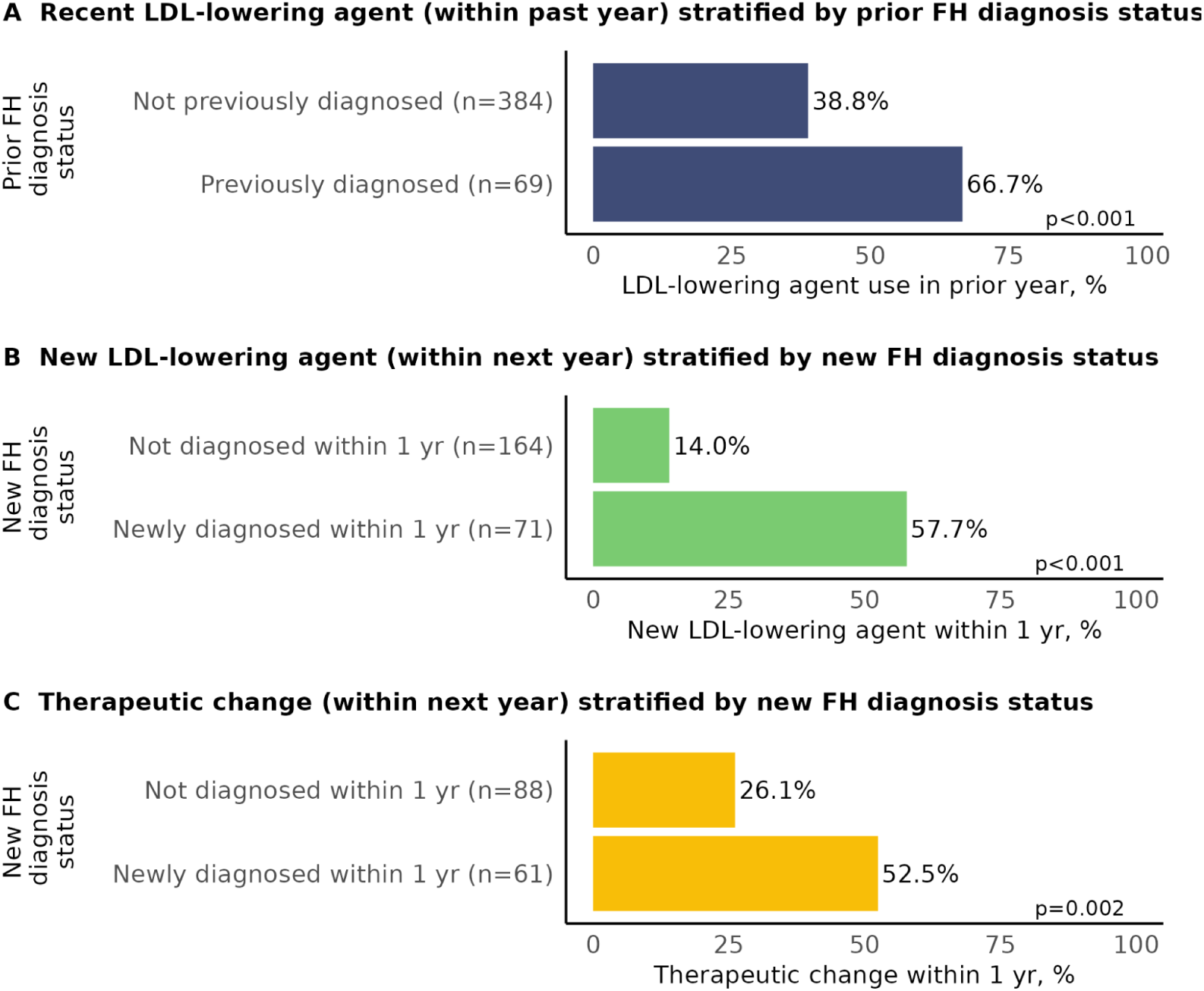
LDL-lowering therapy before and after genetic screening, stratified by FH clinical diagnosis status. FH diagnosis status was defined by the presence or absence of electronic health record documentation of a clinical diagnosis code for FH (SNOMED 398036000 or ICD-10-CM E78.01). Panel A includes all patients. Panel B includes patients without a prior FH diagnosis who had no LDL-lowering agent prescriptions in the prior year. Panel C includes patients without a prior FH diagnosis who had at least one LDL-lowering therapy prescription in the prior year; therapeutic changes include an increase in statin dosage, switching of statin type, or initiation of a statin, ezetimibe, PCSK9 inhibitor, bile acid sequestrant, inclisiran, or bempedoic acid. FH indicates familial hypercholesterolemia; LDL, low-density lipoprotein.

Overall, the most common LDL-lowering agents previously used at any time were statins (97.1% of previously treated patients, including 45.4% with atorvastatin, 42.8% with rosuvastatin, and 7.0% with simvastatin) followed by ezetimibe (38.7%). Among patients with a history of statin use, a majority had most recently used a high-intensity statin (60.5%), followed by moderate-intensity (32.8%) and low-intensity (3.0%) statins. The most recent pre-screening LDL-C result was lower among patients with LDL-lowering therapy in the prior year compared to those without therapy (130 mg/dL vs 156 mg/dL, p<0.0001).

### New documentation of a clinical FH diagnosis and new or modified therapies

Since genetic screening, 177 of 384 patients without a prior diagnosis (46.1%) had a new FH diagnosis subsequently documented in the EHR, after a median of 5.2 months (IQR: 2.6-12.1), whereas 53.9% had no evidence of a new FH diagnosis (after a median total follow-up duration of 25.1 months; IQR: 12.9-56.4). Among all patients, 33% received new or modified LDL-lowering therapy in the first year, and 42% received new/modified therapy within up to 3 years.

Among patients with no LDL-lowering agent use in the prior year, those with a new clinical FH diagnosis code documented in the EHR within the first year post-screening were more likely than those without EHR documentation to be newly prescribed an LDL-lowering agent during the same period (57.7% vs 14.0%, p<0.001; Figure 1B). The most common newly prescribed agents were statins (94.7% of patients, of whom 50.5% received a high-intensity statin and 49.5% received a moderate-intensity statin), ezetimibe (29.3%), and PCSK9 inhibitors (13.3%) (Figure 2A). The median duration to first new therapy was 3.7 months after screening (IQR: 2.2-6.5).

**Figure 2.**
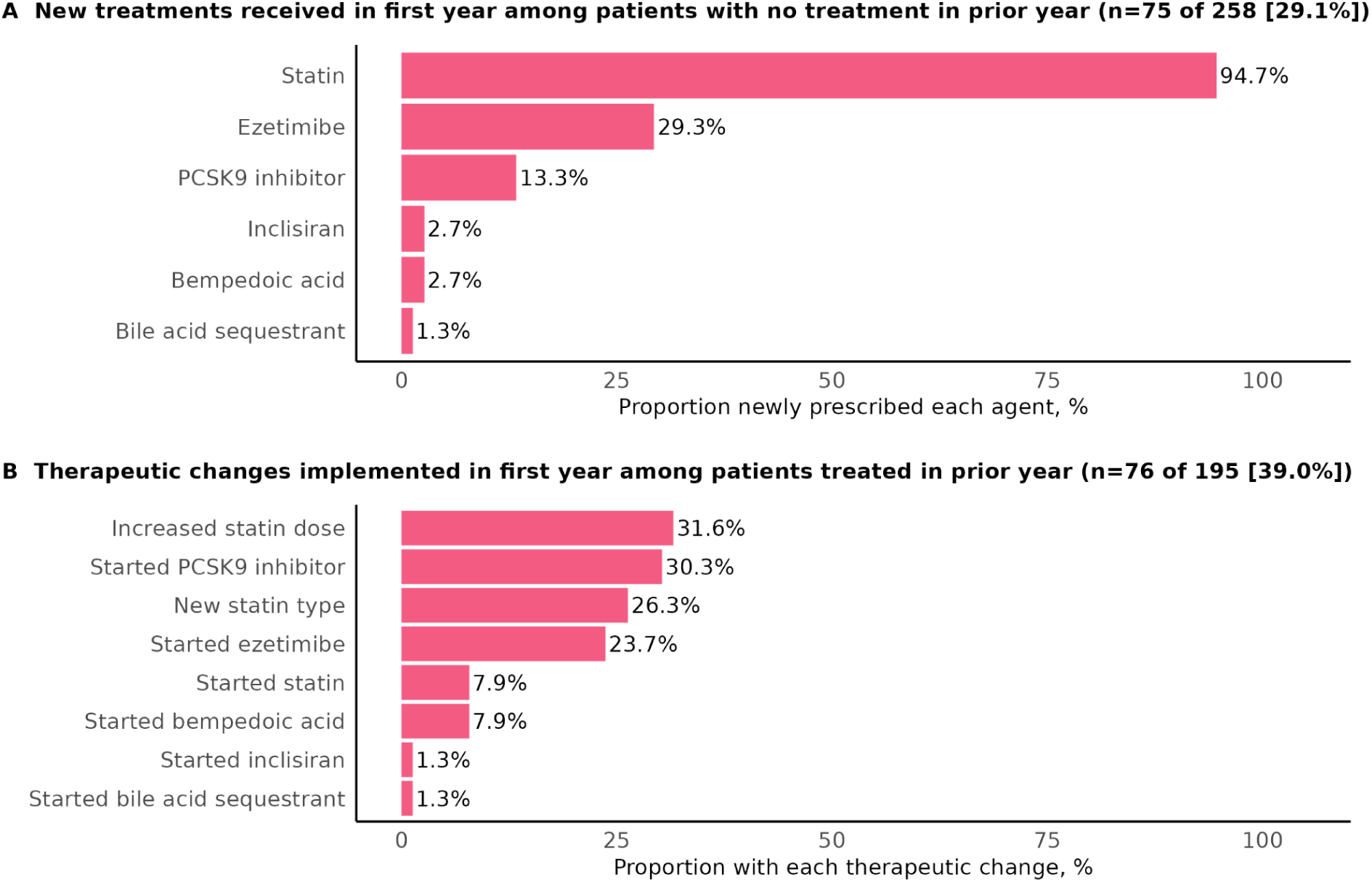
New LDL-lowering agents and therapeutic changes stratified by pre-screening treatment status.

Similarly, among patients with treatment in the prior year, those with the new FH diagnosis code documented were more likely than those without EHR documentation to have a therapeutic change (52.5% vs 26.1%, p=0.002; Figure 1C). The most common therapeutic modifications were increases in statin dosage (31.6%), switches in statin type (26.3%), and starting a PCSK9 inhibitor (30.3%) or ezetimibe (23.7%) (Figure 2B). Differences further stratified by baseline CAD diagnosis status are displayed in Figures S1C-S1F. The median duration to first modified therapy was 5.1 months after screening (IQR: 2.9-7.1).

When comparing against patients who tested negative for FH-associated P/LP variants, rates of new or modified LDL-lowering prescriptions were higher in FH-positive patients post-screening, including when only comparing patients with LDL-C ≥160 mg/dL (Figure 3). This pattern was consistent when restricting the analysis to patients without a prior CAD diagnosis (Figure S2).

**Figure 3.**
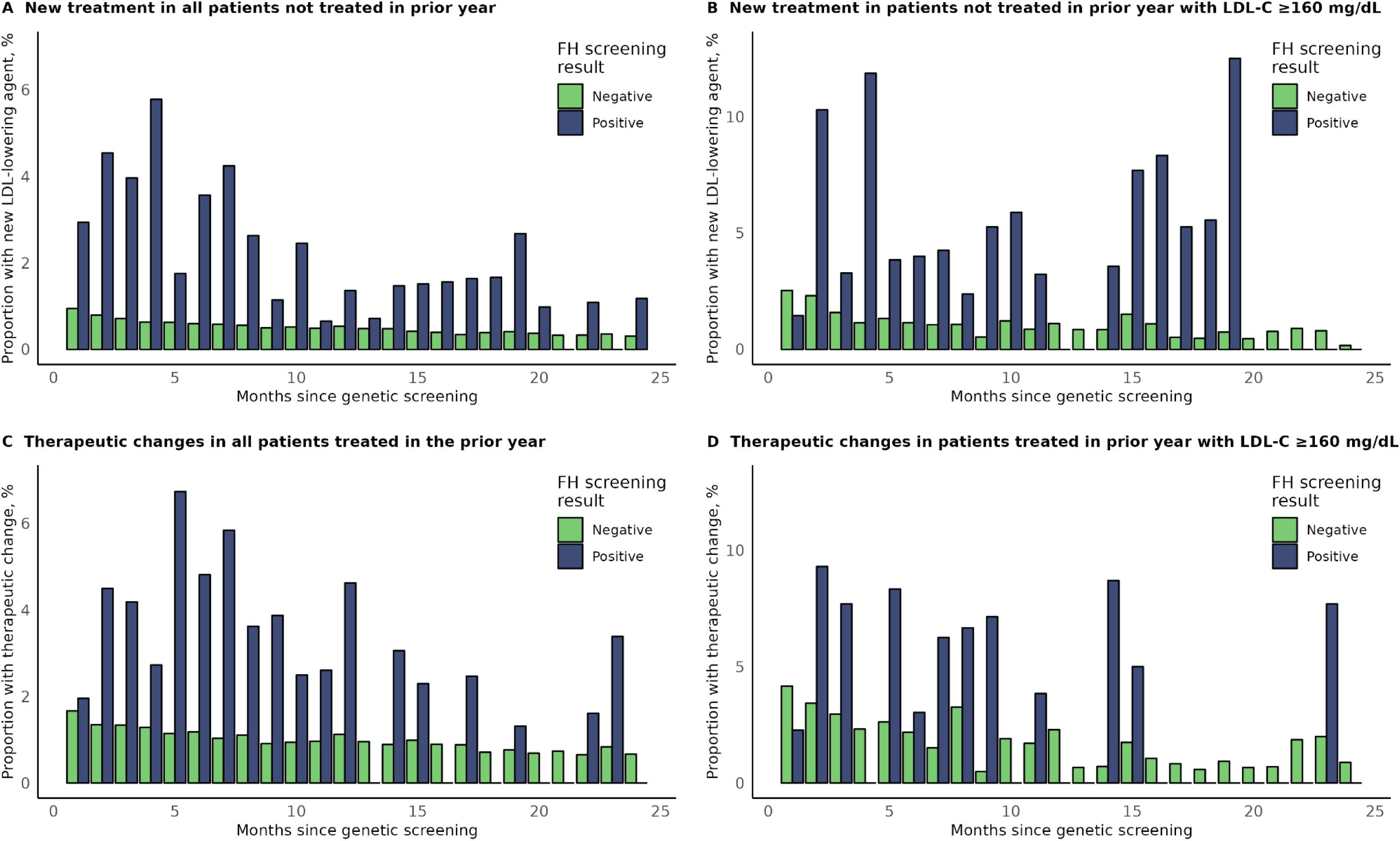
Rates of treatment with new or modified LDL-lowering agents by month since screening among patients who tested positive and negative for FH-associated variants. Proportions of patients with new or modified therapies within each month were calculated among patients with available follow-up for the full month’s duration and without new or modified therapy already having occurred in an earlier month. FH indicates familial hypercholesterolemia; LDL-C, low-density lipoprotein cholesterol

### Changes in serum LDL-C concentrations

Through up to three years post-genetic screening, 339 (74.8%) participants had ≥1 available follow-up LDL-C measurement. During that time, a median of 2 LDL-C measurements were available (IQR: 1-3), with the date of the last measurement having occurred a median of 15.5 months post-screening (IQR: 7.6-23.7). Among those patients, the overall mean maximum reduction in LDL-C was 37 mg/dL, or a 21% reduction from baseline. Overall, patients with new or modified therapies within one year post-screening had a mean LDL-C reduction of 59 mg/dL, compared to 18 mg/dL in patients with no new/modified therapy (difference=41 mg/dL, p<0.001).

Among patients with no LDL-lowering agent use in the year before screening, patients who newly received LDL-lowering treatment within one year post-screening experienced larger maximum LDL-C reductions than those without treatment initiated (mean reductions of 70 mg/dL [39%] vs 14 mg/dL [8%], p<0.001; Figures 4A and 4C). Among patients who had a record of LDL-lowering treatment in the year before screening, patients with a therapeutic change within the next year also had larger reductions than those without a therapeutic change (mean reductions of 53 mg/dL [32%] vs 20 mg/dL [9%], p<0.001; Figures 4B and 4D).

**Figure 4.**
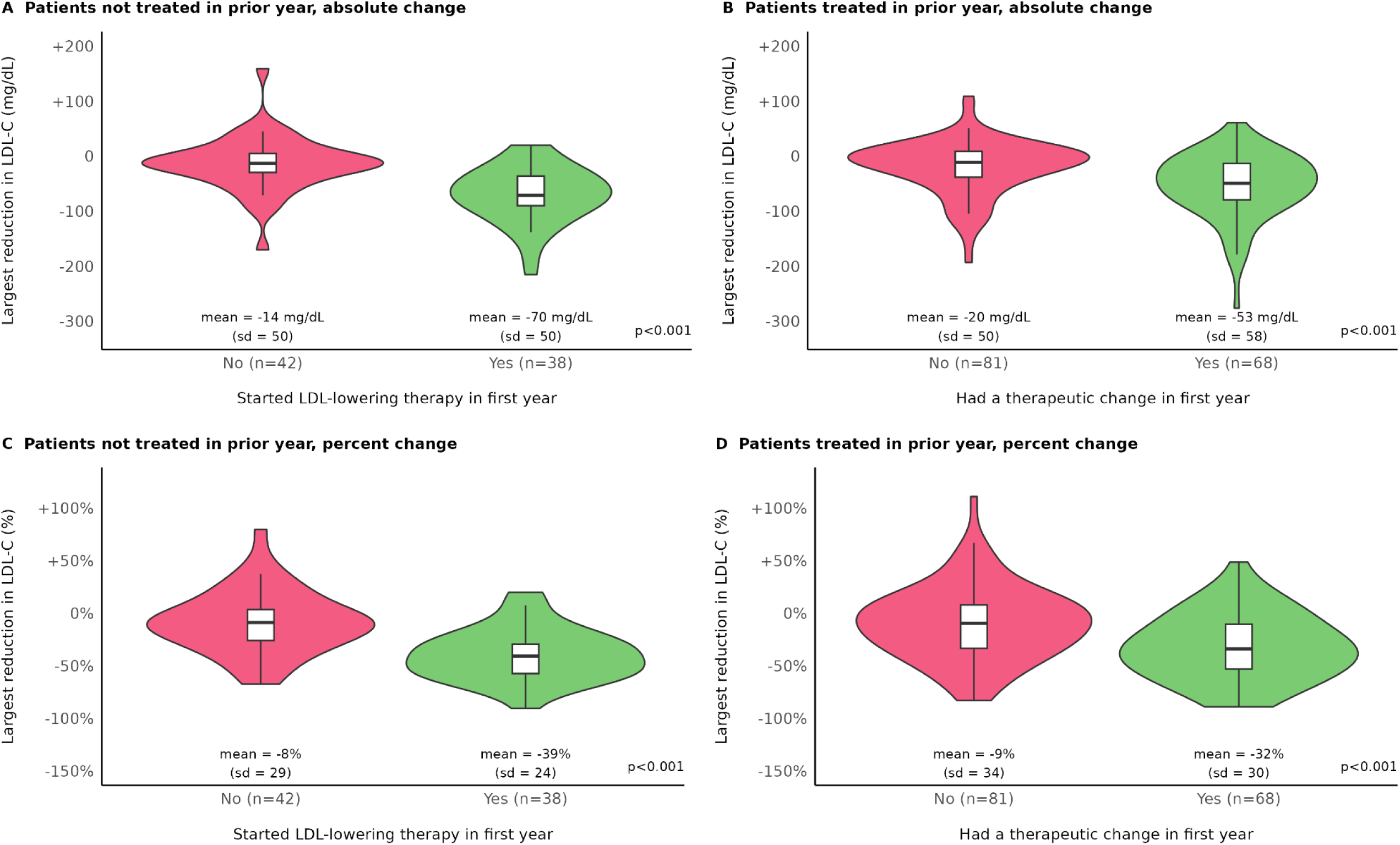
Maximum absolute and percent LDL-C reductions through up to 3 years post-genetic screening, stratified by treatment status. Patients are included if they had a baseline LDL-C result available (through up to 1 year prior to genetic screening) and at least one follow-up LDL-C result. Two patients with their baseline LDL-C testing date within 1-60 days after screening were excluded because their new treatment occurred prior to the baseline LDL-C testing date. Violin plots show the probability density of maximum LDL-C reductions, while overlaid box plots display the median, interquartile range (IQR), and extrema. Whiskers extend to the most extreme data points within 1.5 times the IQR from the box edges. LDL-C indicates low-density lipoprotein cholesterol.

Absolute reductions in LDL-C were larger in patients with higher baseline LDL-C concentrations (Figure 5). However, within each baseline LDL-C category, differences in LDL-C changes largely persisted when comparing patients who did and did not receive new or modified LDL-lowering therapies.

**Figure 5.**
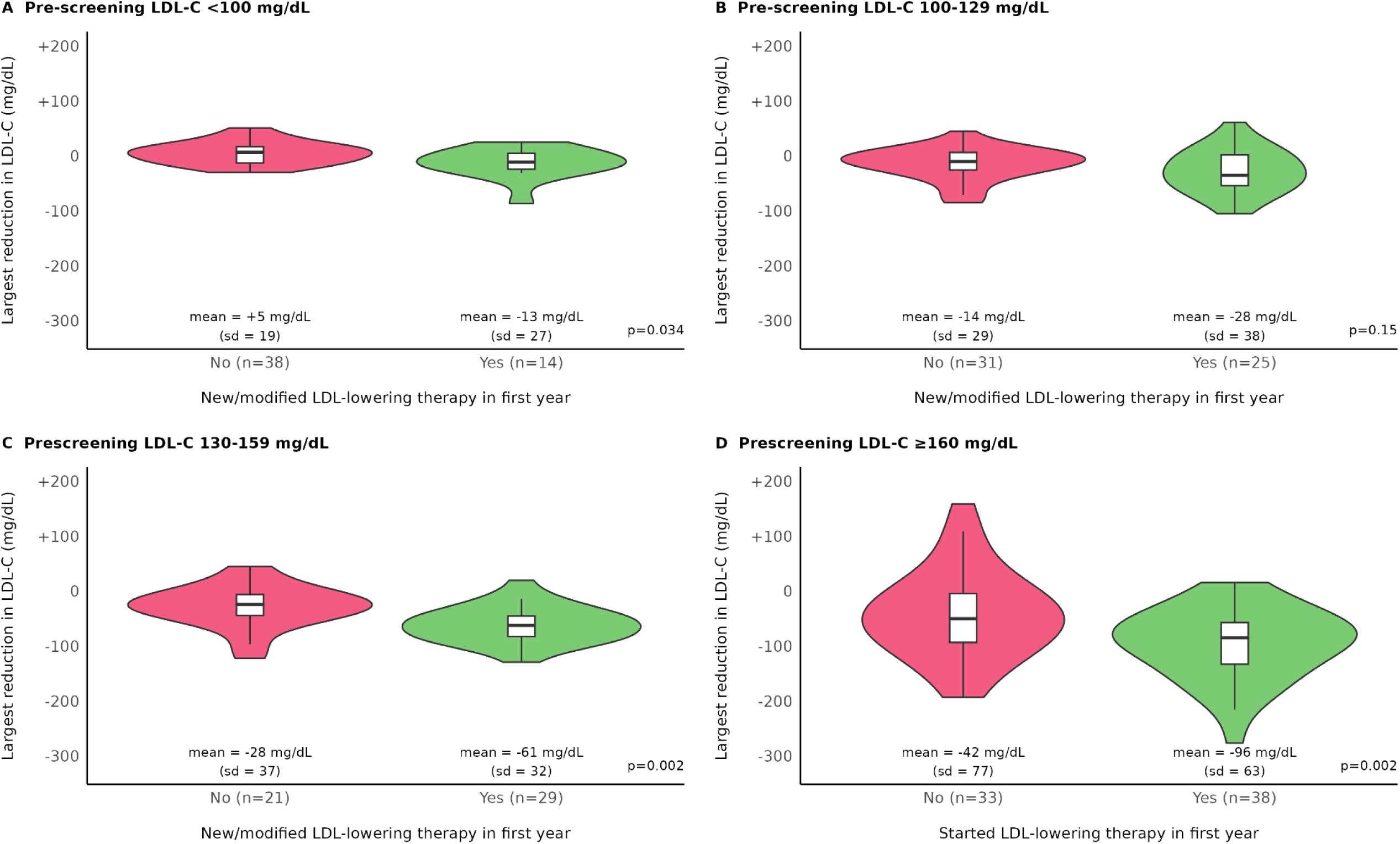
Maximum absolute LDL-C reductions through up to 3 years post-genetic screening, stratified by pre-screening LDL-C concentration. Patients are included if they had a baseline LDL-C result available (through up to 1 year prior to genetic screening) and at least one follow-up LDL-C result. Two patients with their baseline LDL-C testing date within 1-60 days after screening were excluded because their new treatment occurred prior to the baseline LDL-C testing date.Violin plots show the probability density of maximum LDL-C reductions, while overlaid box plots display the median, interquartile range (IQR), and extrema. Whiskers extend to the most extreme data points within 1.5 times the IQR from the box edges. LDL-C indicates low-density lipoprotein cholesterol.

Individual longitudinal trajectories for changes in LDL-C are displayed in Figure S3. In patients with therapeutic additions or changes, LDL-C reductions were largely persistent throughout the follow-up period. Although average reductions were lower in patients without therapeutic additions/changes, a subset of those patients similarly had persistent reductions.

Among patients with baseline LDL-C ≥70 mg/dL, those with new or modified therapies were also more likely than those without therapeutic additions or changes to achieve either a ≥50% reduction in LDL-C or LDL-C <70 mg/dL (42.0% vs 15.3%, p<0.001) (Figure 6). The median time to achievement of this target goal was 8.9 months (IQR: 4.9-13.3), with a median of 50% of all post-screening measurements being below the threshold in this group (IQR: 33%-100%).

**Figure 6.**
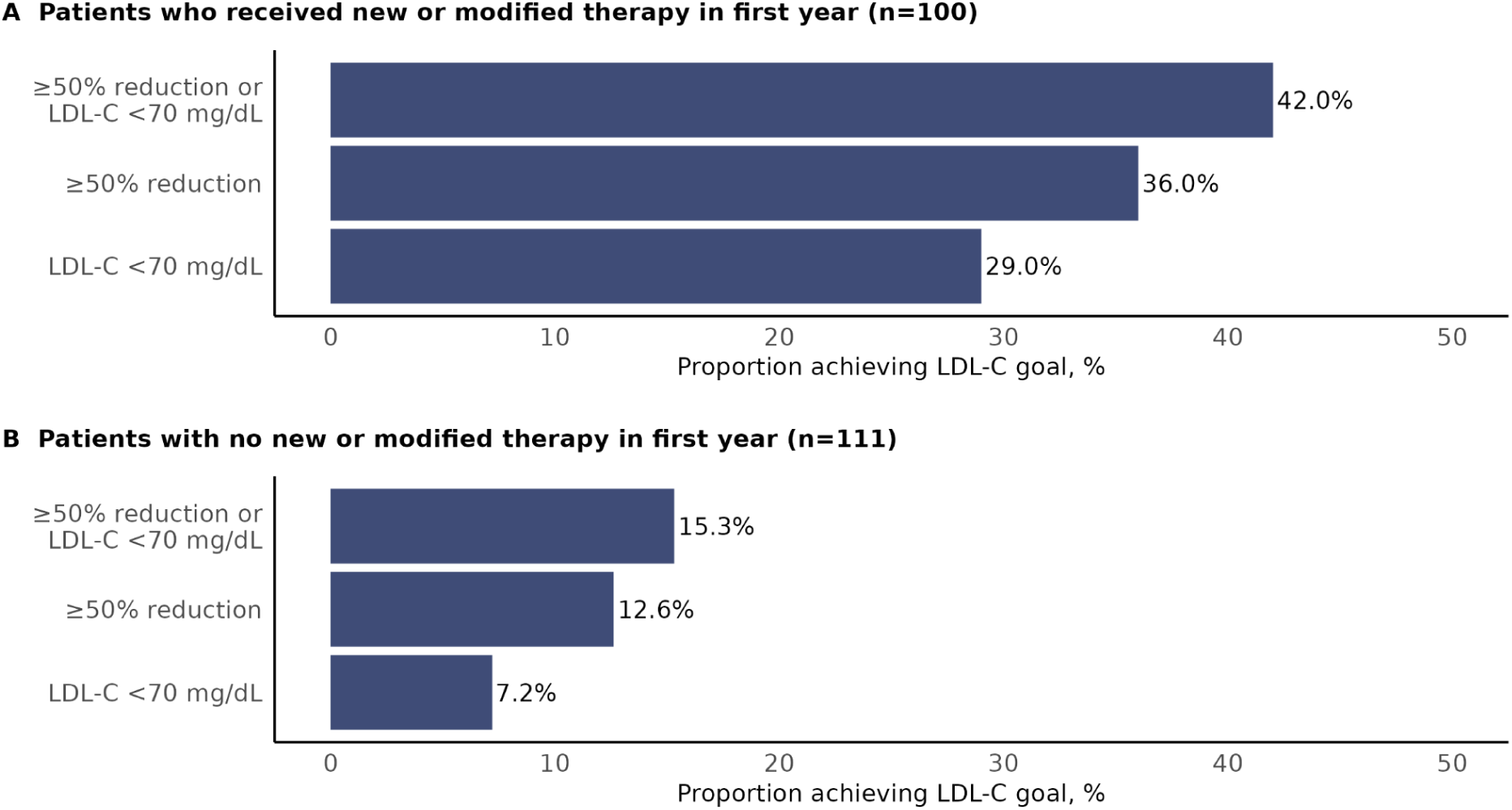
Achievement of LDL-C target goals through up to 3 years post-genetic screening, stratified by treatment status. Patients are included if they had a baseline LDL-C result available (through up to 1 year prior to genetic screening) and at least one follow-up LDL-C result. 18 patients with pre-screening LDL-C <70 mg/dL were excluded from this figure. One patient with the baseline LDL-C testing date within 1-60 days after screening was excluded because the new treatment occurred prior to the baseline LDL-C testing date. P<0.001 for all comparisons between patients with vs without new/modified therapy. LDL-C indicates low-density lipoprotein cholesterol.

Overall, patients with a prior CAD diagnosis were more likely than those without a CAD diagnosis to achieve either a ≥50% reduction in LDL-C or LDL-C <70 mg/dL (47.2% vs 24.0%, p=0.009). This trend was similarly observed among individuals with new or modified therapies (62.5% vs 38.1%, p=0.12) and those without new or modified therapies (35.0% vs 11.0%, p=0.013) (Figure S4).

## Discussion

Within a large-scale population genomics program, we demonstrate the powerful potential of population screening for improving lipid management in individuals with FH-associated pathogenic variants. Our study reinforces the significant underdiagnosis of FH in standard clinical practice, as fewer than 1 in 6 patients with an FH-associated variant had a prior clinical FH diagnosis, despite more than two-thirds having been previously diagnosed with general hyperlipidemia. Following disclosure of positive FH screening results, we observed a significant shift in clinical management. Many patients received new or modified lipid-lowering therapies within the subsequent year, with higher rates of therapeutic changes among FH-positive patients compared to FH-negative counterparts. Notably, despite nearly two-thirds of FH-positive patients having a history of LDL-lowering therapy, therapeutic modifications post-screening were associated with substantial LDL-C reductions. The mean reduction in

LDL-C among FH-positive patients who experienced therapeutic changes was 59 mg/dL, which approximately corresponds to a 32% reduction in risk of cardiovascular events.^19^ These findings emphasize the importance of accurate FH diagnosis for optimal downstream clinical management.

Because FH is characterized by lifelong high LDL-C levels and subsequent elevated risk of premature cardiovascular events, LDL-C targets are more stringent for individuals with FH than in the general population.^6,7^ Among patients with pre-screening LDL-C ≥70 mg/dL who experienced a therapeutic change post-screening, 42% subsequently achieved an LDL-C target of either <70 mg/dL or at least a 50% reduction within up to three years of available follow-up, demonstrating the potential impact of genetic risk identification on lipid control. The proportion achieving this target was lower in those with no new/modified therapies, and this trend was observed among patients with and without a prior CAD diagnosis. However, gaps remain in achieving optimal control for all individuals. While our findings demonstrate that genomic screening can drive more effective intervention, further efforts are needed to close these gaps and optimize long-term treatment strategies.

Surprisingly, only slightly more than half of the patients with an FH-associated variant had subsequent EHR documentation of a clinical diagnostic code for FH (e.g., ICD-10-CM code E78.01 in the problem list), despite all patients having had one or more medical visits at least six months after genetic screening. While the FH diagnosis code was used across all participating health systems, it is possible that some patients had their FH diagnosis documented in other structured (e.g., genomic testing indicator) or unstructured (e.g., clinical notes) EHR fields that were not available for analysis.

Furthermore, lipid management changes were significantly more common in individuals with new EHR documentation of the FH diagnosis code compared to those without—twice as likely among patients already treated in the prior year and four times as likely among patients not treated in the prior year. While this association is not necessarily causal, it suggests that formal recognition of FH within the medical record may reflect, or facilitate, more proactive lipid management, including the initiation or intensification of therapy. Previous studies have also demonstrated improved compliance with lipid-lowering medication among patients with a genetically confirmed FH diagnosis.^20^ Beyond identifying FH-associated variants through genetic screening, it is important that this information be effectively integrated into clinical workflows to drive appropriate therapeutic action. Possible approaches include the development of standardized scripting for new FH diagnoses, automated scheduling with a genetic counselor, creation of a health care maintenance topic driven off of the result (e.g., reminding clinicians to order LDL-C and refer to a lipid clinic), and generation of a genomics indicator viewable to both the patient and provider, adding broader visibility to the diagnosis. Moreover, the identification of FH cases through genetic screening enables the implementation of cascade testing among family members.^21^ This approach can amplify the impact of each FH diagnosis, potentially identifying multiple additional cases within families, enabling early intervention and extending the benefits of population screening.

Even in FH patients with no therapeutic changes, many experienced persistent reductions in LDL-C levels, which may be related to other management changes. This could include behavioral modifications (e.g., diet, exercise), increased healthcare engagement and LDL-C monitoring, improved medication adherence, weight loss, or reductions in systemic inflammation. Although not expected to be common, it is possible that some patients obtained lipid-lowering therapy from another health system, which could have resulted in underascertainment of lipid-lowering agent records.

Additionally, our study relied on prescription records to define LDL-lowering treatment status and it was unknown whether patients were fully compliant with prescribed treatment regimens. Factors such as variations in laboratory measurements or spontaneous fluctuations in lipid levels over time may have contributed to measurement error. For this analysis, data on fasting status was not available; however, non-fasting status is less likely to affect LDL-C levels, compared to triglyceride levels. Heterogeneity in standard care procedures, clinical workflows, provider practices, and patient populations across and within participating health systems may have contributed to observed differences in outcomes. While these factors could not be fully characterized in the current study, they represent important considerations in planning future implementations of genomic screening programs.

In summary, population genomic screening for FH demonstrated significant potential for cardiovascular disease prevention, yielding lipid management improvements and substantial LDL-C reductions. The more pronounced impact in patients with newly documented FH diagnoses highlights the critical need to integrate genomic results into clinical workflows. Future efforts should focus on optimizing result documentation, enhancing long-term lipid management, and evaluating cardiovascular outcomes to maximize the benefits of genomic screening.

## Data Availability

The Helix Research Network (HRN) data are available to qualified researchers upon reasonable request and with permission of the HRN Steering Committee and Helix. Researchers who would like to obtain the raw genotype data related to this study will be presented with a Data Use Agreement which requires that participants will not be re-identified and that no data will be shared between individuals, third parties, or uploaded onto public domains. The HRN encourages collaboration with scientific researchers on an individual basis. Examples of restrictions that will be considered in requests to access data include but are not limited to: 1) whether the request comes from an academic institution in good standing and will collaborate with our team to protect the privacy of the participants and the security of the data requested; 2) type and amount of data requested; 3) feasibility of the research suggested; and 4) amount of resource allocation for Helix and HRN member institutions required to support a collaboration.

## Acknowledgments

We acknowledge the Helix bioinformatics and lab teams for their contributions to the production of the exome sequencing pipeline as well as the research administration team for coordinating the project. We thank all of the participants of DNA Answers, GeneConnect, the Genetic Insights Project, the Healthy Nevada Project, ImagineYou, In Our DNA SC, myGenetics, and The Gene Health Project.

## Supplemental Material

Supplemental Data Files 1-2

Figures S1-S4

## Supplemental Material

**Supplemental Data File 1. List of evaluated pathogenic variants in *LDLR*, *APOB*, and *PCSK9* genes associated with familial hypercholesterolemia.** Supplemental data file: Supplemental_Data_File_1.csv.

**Supplemental Data File 2. Observational Medical Outcomes Partnership (OMOP) Common Data Model (CDM) concept sets used to define LDL-lowering agents and electronic health record-based diagnosis measures.** Supplemental data file: Supplemental_Data_File_2.csv.

**Figure S1.**
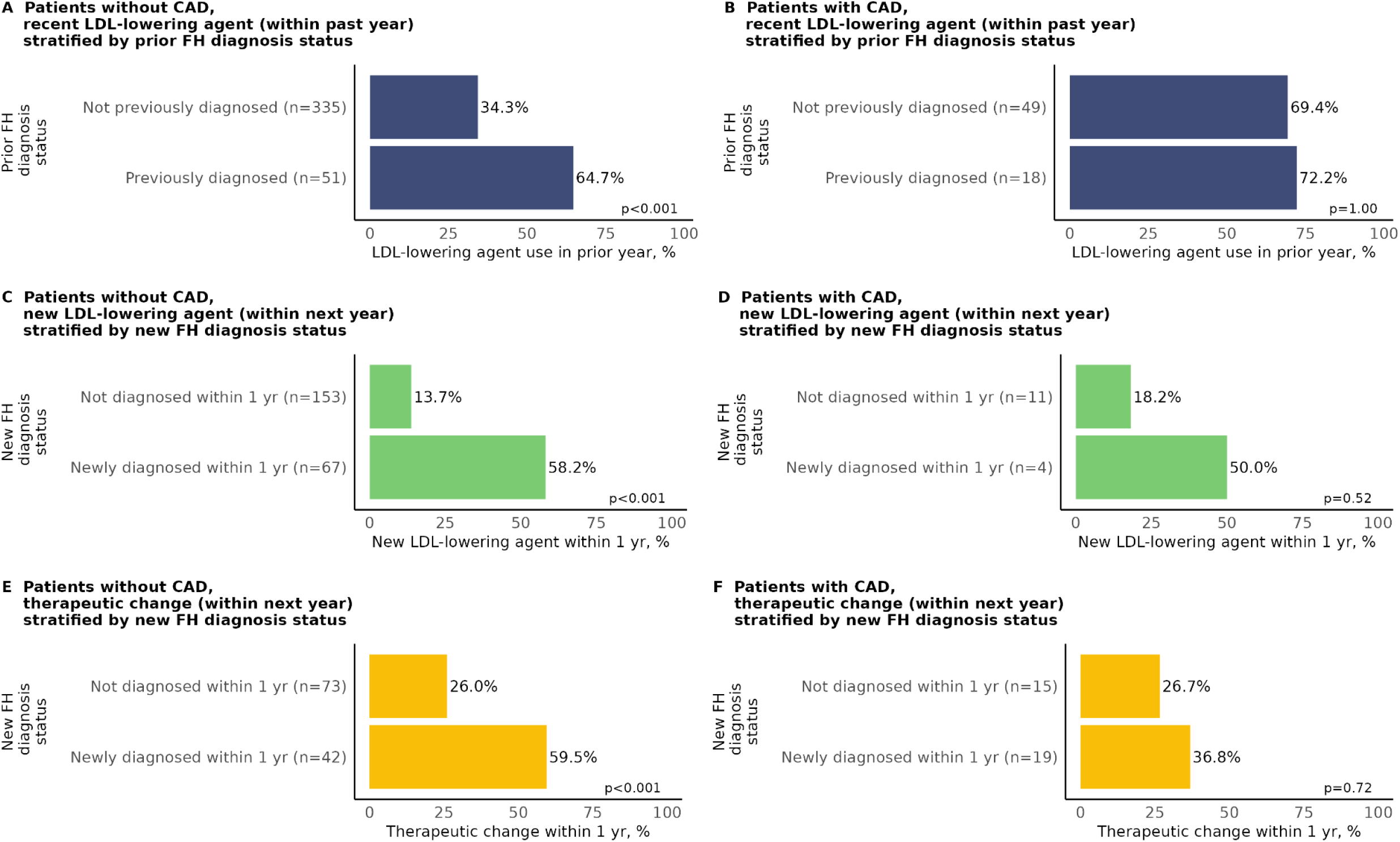
LDL-lowering therapy before and after genetic screening, stratified by coronary artery disease status and FH clinical diagnosis status. FH diagnosis status was defined by the presence or absence of electronic health record documentation of a clinical diagnosis code for FH (SNOMED 398036000 or ICD-10-CM E78.01). Panels A-B includes all patients without and with CAD, respectively. Panels C-D includes those patients without a prior FH diagnosis who had no LDL-lowering agent prescriptions in the prior year. Panels E-F include those patients without a prior FH diagnosis who had at least one LDL-lowering therapy prescription in the prior year; therapeutic changes include an increase in statin dosage, switching of statin type, or initiation of a statin, ezetimibe, PCSK9 inhibitor, bile acid sequestrant, inclisiran, or bempedoic acid. CAD indicates coronary artery disease; FH, familial hypercholesterolemia.

**Figure S2.**
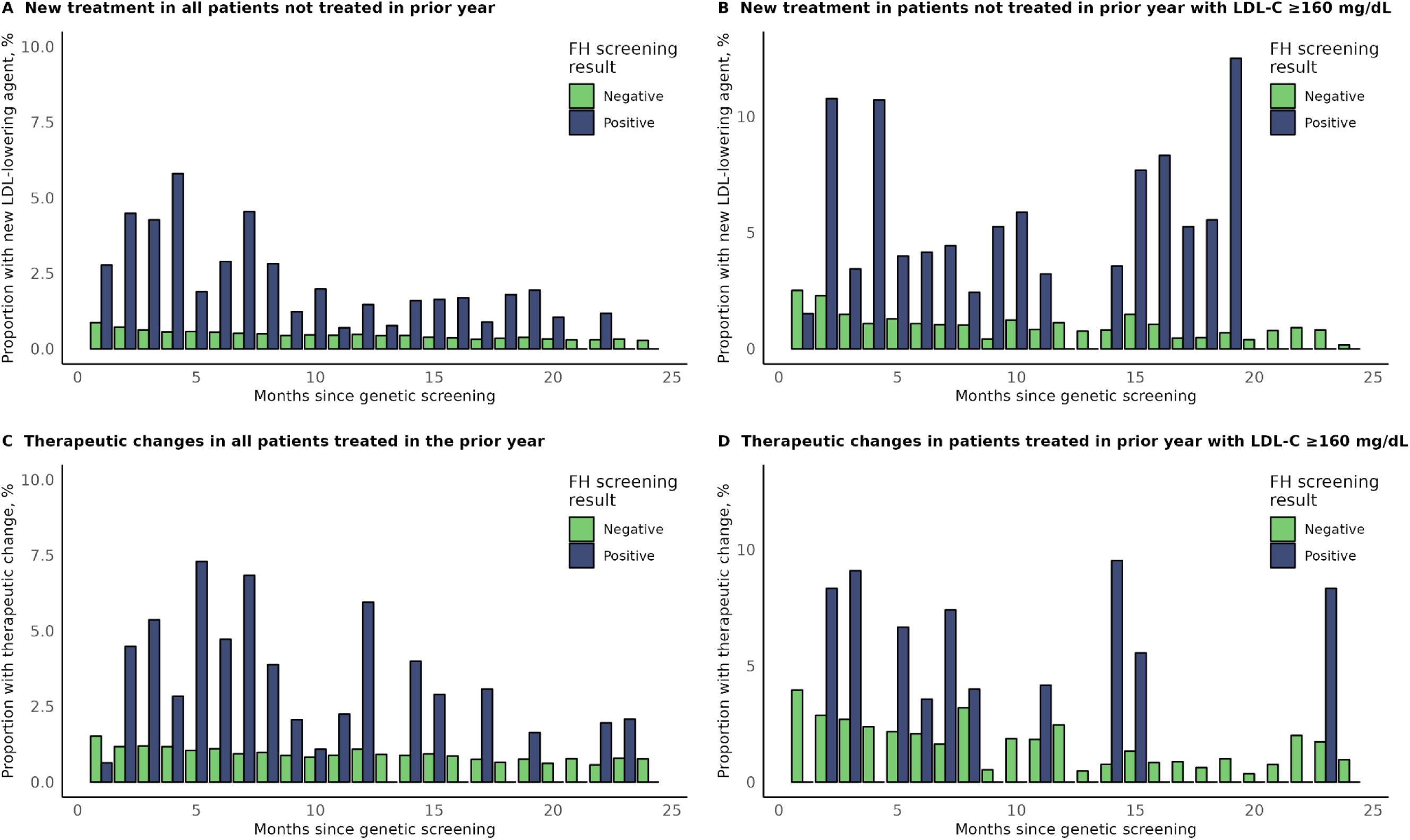
Rates of treatment with new or modified LDL-lowering agents by month since screening among patients without a coronary artery disease diagnosis who tested positive and negative for FH-associated variants. Proportions of patients with new or modified therapies within each month were calculated among patients with available follow-up for the full month’s duration and without new or modified therapy already having occurred in an earlier month. FH indicates familial hypercholesterolemia.

**Figure S3.**
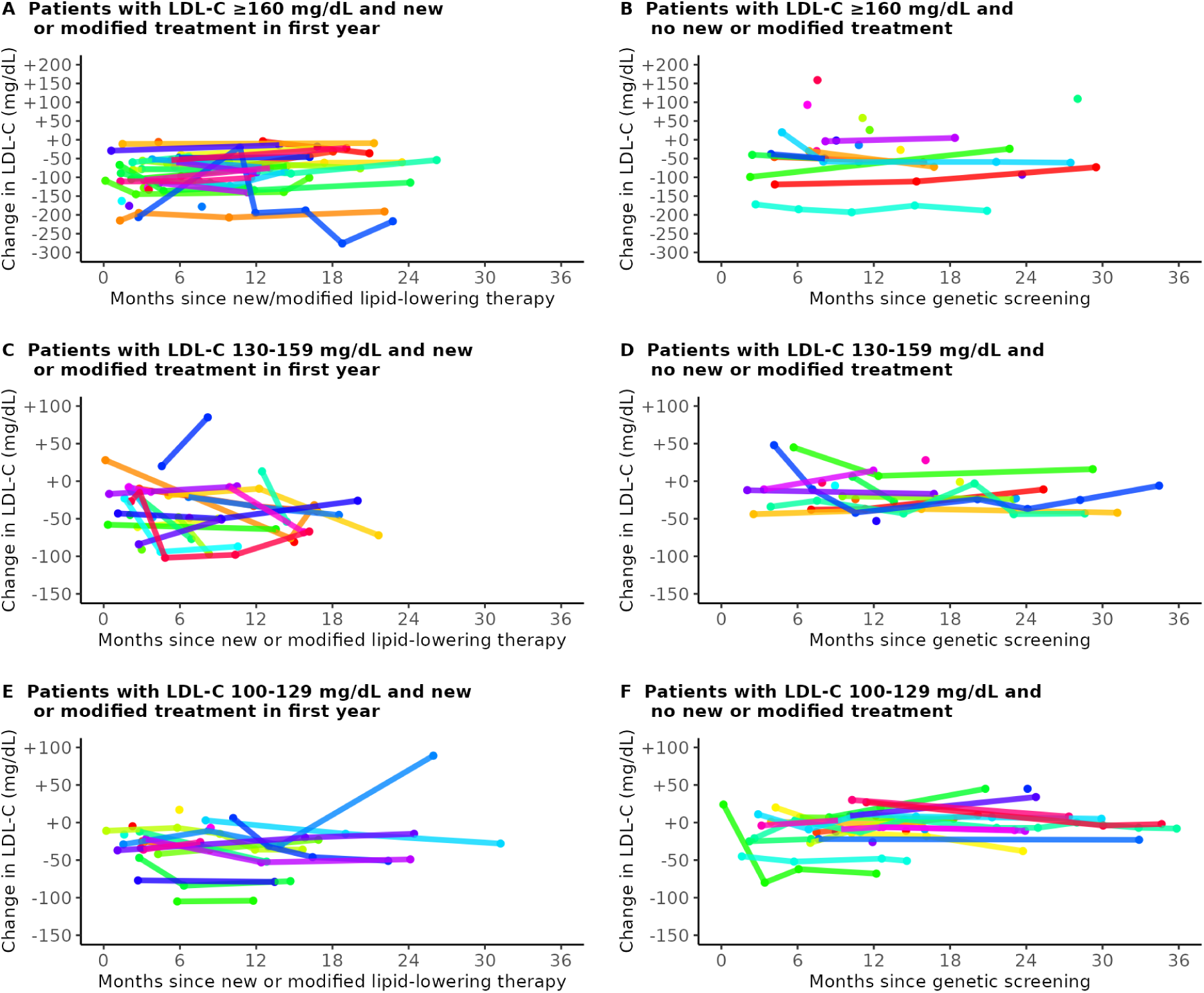
Trajectories of LDL-C changes through up to 3 years after screening, stratified by baseline LDL-C concentration and lipid-lowering agent treatment status. Includes patients with at least 1 LDL-C result spanning at least 6 months after genetic screening (for patients with no new/modified therapy) or after receipt of new/modified therapy (for those with new/modified therapy), in the first 3 years since screening. In those with no therapeutic addition/change (through up to 3 years), the y-axis is time since screening. In those with a therapeutic addition/change, the y-axis is time since first new/modified lipid-lowering therapy in the first year.

**Figure S4.**
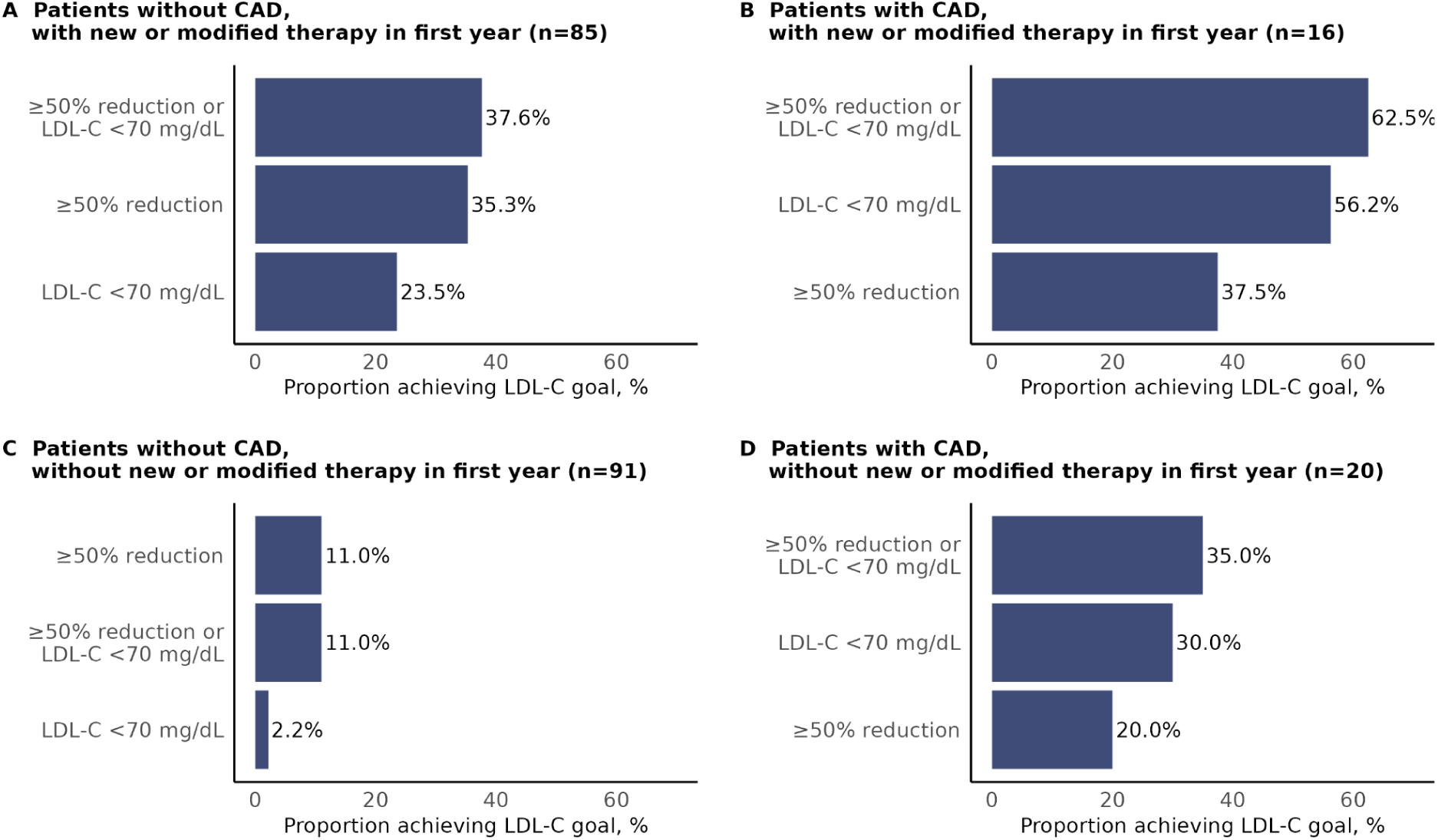
Achievement of LDL-C target goals through up to 3 years post-genetic screening, stratified by CAD diagnosis status and treatment status. Patients are included if they had a baseline LDL-C result available (through up to 1 year prior to genetic screening) and at least one follow-up LDL-C result. 18 patients with pre-screening LDL-C <70 mg/dL were excluded from this figure. One patient with the baseline LDL-C testing date within 1-60 days after screening was excluded because the new treatment occurred prior to the baseline LDL-C testing date. Among patients without a CAD diagnosis, p<0.001 for each of the three comparisons between patients with vs without new/modified therapy. Among patients with a CAD diagnosis, p=0.19, p=0.29, and p=0.21 for each of the three comparisons between patients with vs without new/modified therapy. CAD indicates coronary artery disease.

## References

1. McGowan MP, Hosseini Dehkordi SH, Moriarty PM, Duell PB. Diagnosis and treatment of heterozygous familial hypercholesterolemia. J Am Heart Assoc 2019;8:e013225.

2. Sturm AC, Knowles JW, Gidding SS, Ahmad ZS, Ahmed CD, Ballantyne CM, et al. Clinical genetic testing for familial hypercholesterolemia: JACC scientific expert panel. J Am Coll Cardiol 2018;72:662–680.

3. Soutar AK, Naoumova RP. Mechanisms of disease: genetic causes of familial hypercholesterolemia. Nat Clin Pract Cardiovasc Med 2007;4:214–225.

4. Ison HE, Clarke SL, Knowles JW. Familial hypercholesterolemia. GeneReviews(®), Seattle (WA): University of Washington, Seattle; 1993.

5. Nordestgaard BG, Chapman MJ, Humphries SE, Ginsberg HN, Masana L, Descamps OS, et al. Familial hypercholesterolaemia is underdiagnosed and undertreated in the general population: guidance for clinicians to prevent coronary heart disease: consensus statement of the European Atherosclerosis Society. Eur Heart J 2013;34:3478–3490a.

6. Grundy SM, Stone NJ, Bailey AL, Beam C, Birtcher KK, Blumenthal RS, et al. 2018 AHA/ACC/AACVPR/AAPA/ABC/ACPM/ADA/AGS/APhA/ASPC/NLA/PCNA guideline on the management of blood cholesterol: a report of the American College of Cardiology/American Heart Association Task Force on Clinical Practice Guidelines. J Am Coll Cardiol 2019;73:e285–e350.

7. Authors/Task Force Members, ESC Committee for Practice Guidelines (CPG), ESC National Cardiac Societies. 2019 ESC/EAS guidelines for the management of dyslipidaemias: Lipid modification to reduce cardiovascular risk. Atherosclerosis 2019;290:140–205.

8. Stark Z, Dolman L, Manolio TA, Ozenberger B, Hill SL, Caulfied MJ, et al. Integrating genomics into healthcare: A global responsibility. Am J Hum Genet 2019;104:13–20.

9. Buchanan AH, Lester Kirchner H, Schwartz MLB, Kelly MA, Schmidlen T, Jones LK, et al. Clinical outcomes of a genomic screening program for actionable genetic conditions. Genet Med 2020;22:1874–1882.

10. Grzymski JJ, Elhanan G, Morales Rosado JA, Smith E, Schlauch KA, Read R, et al. Population genetic screening efficiently identifies carriers of autosomal dominant diseases. Nat Med 2020;26:1235–1239.

11. Jones LK, Chen N, Hassen DA, Betts MN, Klinger T, Hartzel DN, et al. Impact of a population genomic screening program on health behaviors related to familial hypercholesterolemia risk reduction. Circ Genom Precis Med 2022;15:e003549.

12. Bellows BK, Khera AV, Zhang Y, Ruiz-Negrón N, Stoddard HM, Wong JB, et al. Estimated yield of screening for Heterozygous familial hypercholesterolemia with and without genetic testing in US adults. J Am Heart Assoc 2022;11:e025192.

13. Simon Broome Register Group. Risk of fatal coronary heart disease in familial hypercholesterolaemia. BMJ 1991;303:893–896.

14. WHO Human Genetics Programme. Familial hypercholesterolaemia (FH) : report of a second WHO consultation, Geneva, 4 September 1998. World Health Organization; 1999.

15. Helix Research Network (HRN). ClinicalTrialsGov 2024. https://clinicaltrials.gov/study/NCT06057181 (accessed March 13, 2025).

16. Centers for Disease Control and Prevention. Tier 1 genomic applications toolkit for public health departments 2014. https://archive.cdc.gov/#/details?url=https://www.cdc.gov/genomics/implementation/toolkit/index.htm (accessed February 12, 2025).

17. Cirulli ET, White S, Read RW, Elhanan G, Metcalf WJ, Tanudjaja F, et al. Genome-wide rare variant analysis for thousands of phenotypes in over 70,000 exomes from two cohorts. Nat Commun 2020;11:542.

18. Voss EA, Makadia R, Matcho A, Ma Q, Knoll C, Schuemie M, et al. Feasibility and utility of applications of the common data model to multiple, disparate observational health databases. J Am Med Inform Assoc 2015;22:553–564.

19. Cholesterol Treatment Trialists’ (CTT) Collaboration, Baigent C, Blackwell L, Emberson J, Holland LE, Reith C, et al. Efficacy and safety of more intensive lowering of LDL cholesterol: a meta-analysis of data from 170,000 participants in 26 randomised trials. Lancet 2010;376:1670–1681.

20. Umans-Eckenhausen MAW, Defesche JC, van Dam MJ, Kastelein JJP. Long-term compliance with lipid-lowering medication after genetic screening for familial hypercholesterolemia. Arch Intern Med 2003;163:65–68.

21. Morris JK, Wald DS, Wald NJ. The evaluation of cascade testing for familial hypercholesterolemia. Am J Med Genet A 2012;158A:78–84.

