## Supplemental Material for "Population genomic screening leads to improved lipid management in patients with familial hypercholesterolemia"

**Supplemental Data File 1. List of evaluated pathogenic variants in *LDLR*, *APOB*, and *PCSK9* genes associated with familial hypercholesterolemia.** Supplemental data file: Supplemental_Data_File_1.csv.

**Supplemental Data File 2. Observational Medical Outcomes Partnership (OMOP) Common Data Model (CDM) concept sets used to define LDL-lowering agents and electronic health record-based diagnosis measures.** Supplemental data file: Supplemental_Data_File_2.csv.

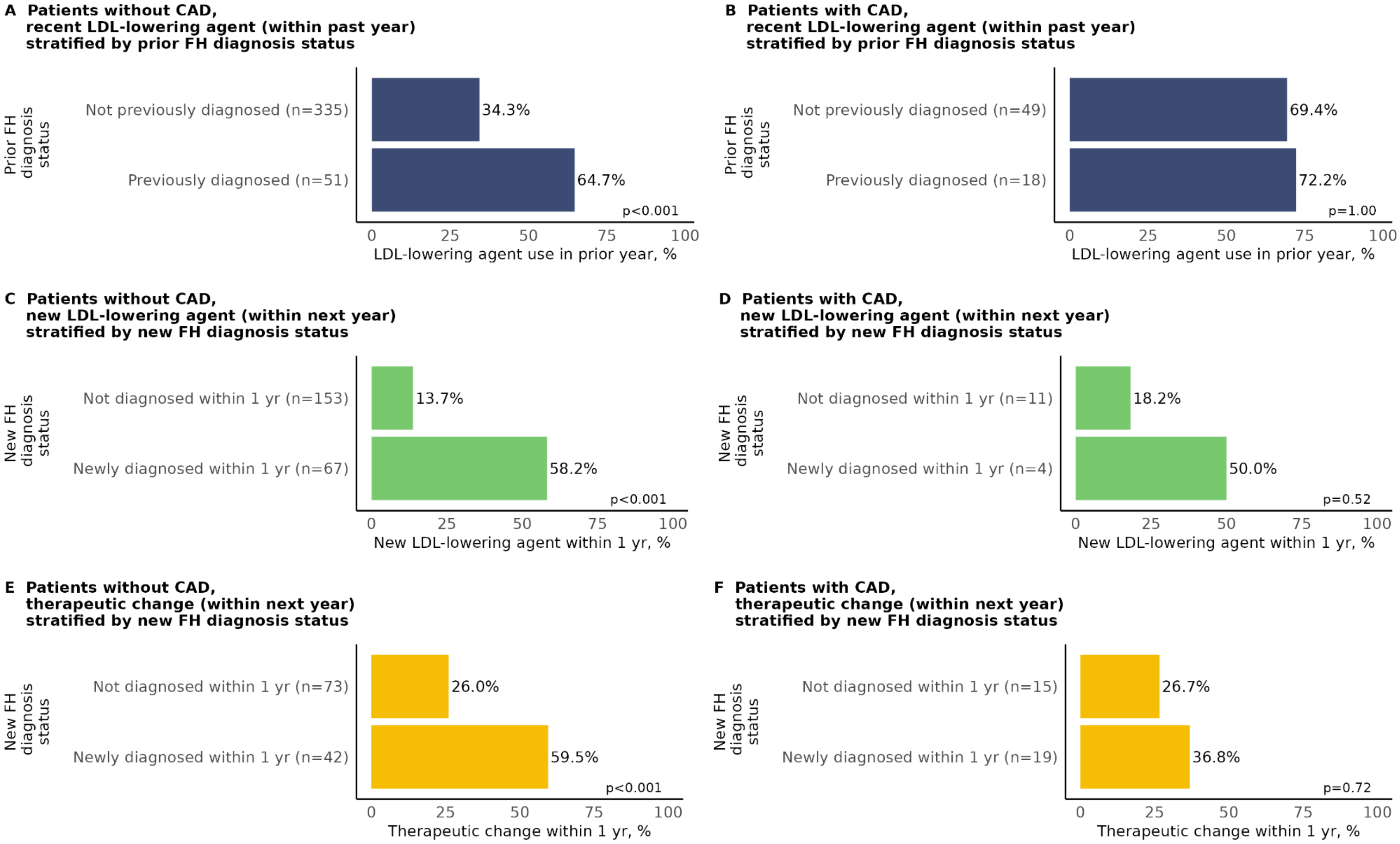


**Figure S1. LDL-lowering therapy before and after genetic screening, stratified by coronary artery disease status and FH clinical diagnosis status.** FH diagnosis status was defined by the presence or absence of electronic health record documentation of a clinical diagnosis code for FH (SNOMED 398036000 or ICD-10-CM E78.01). Panels A-B includes all patients without and with CAD, respectively. Panels C-D includes those patients without a prior FH diagnosis who had no LDL-lowering agent prescriptions in the prior year. Panels E-F include those patients without a prior FH diagnosis who had at least one LDL-lowering therapy prescription in the prior year; therapeutic changes include an increase in statin dosage, switching of statin type, or initiation of a statin, ezetimibe, PCSK9 inhibitor, bile acid sequestrant, inclisiran, or bempedoic acid. CAD indicates coronary artery disease; FH, familial hypercholesterolemia.

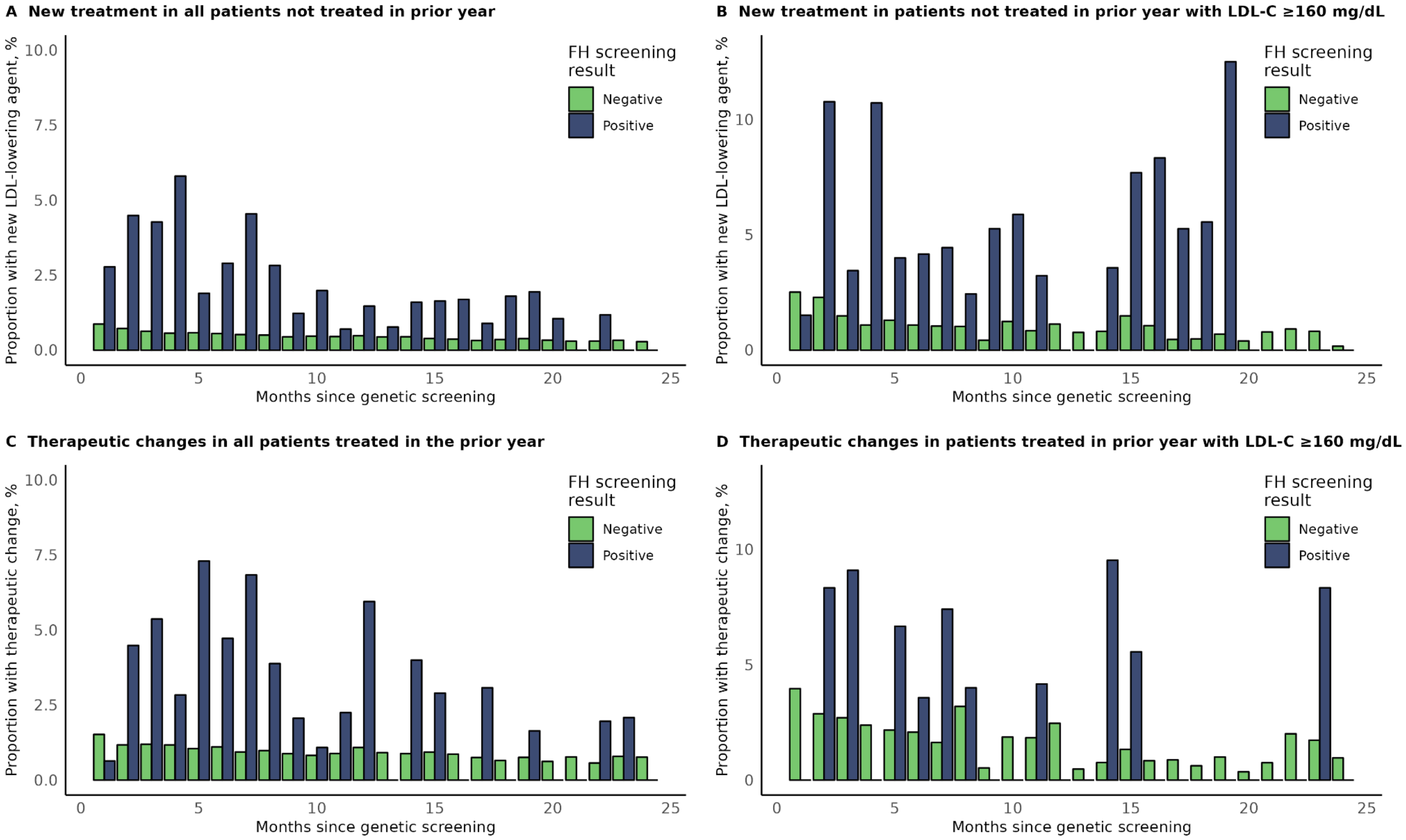


**Figure S2. Rates of treatment with new or modified LDL-lowering agents by month since screening among patients without a coronary artery disease diagnosis who tested positive and negative for FH-associated variants.** Proportions of patients with new or modified therapies within each month were calculated among patients with available follow-up for the full month's duration and without new or modified therapy already having occurred in an earlier month. FH indicates familial hypercholesterolemia.

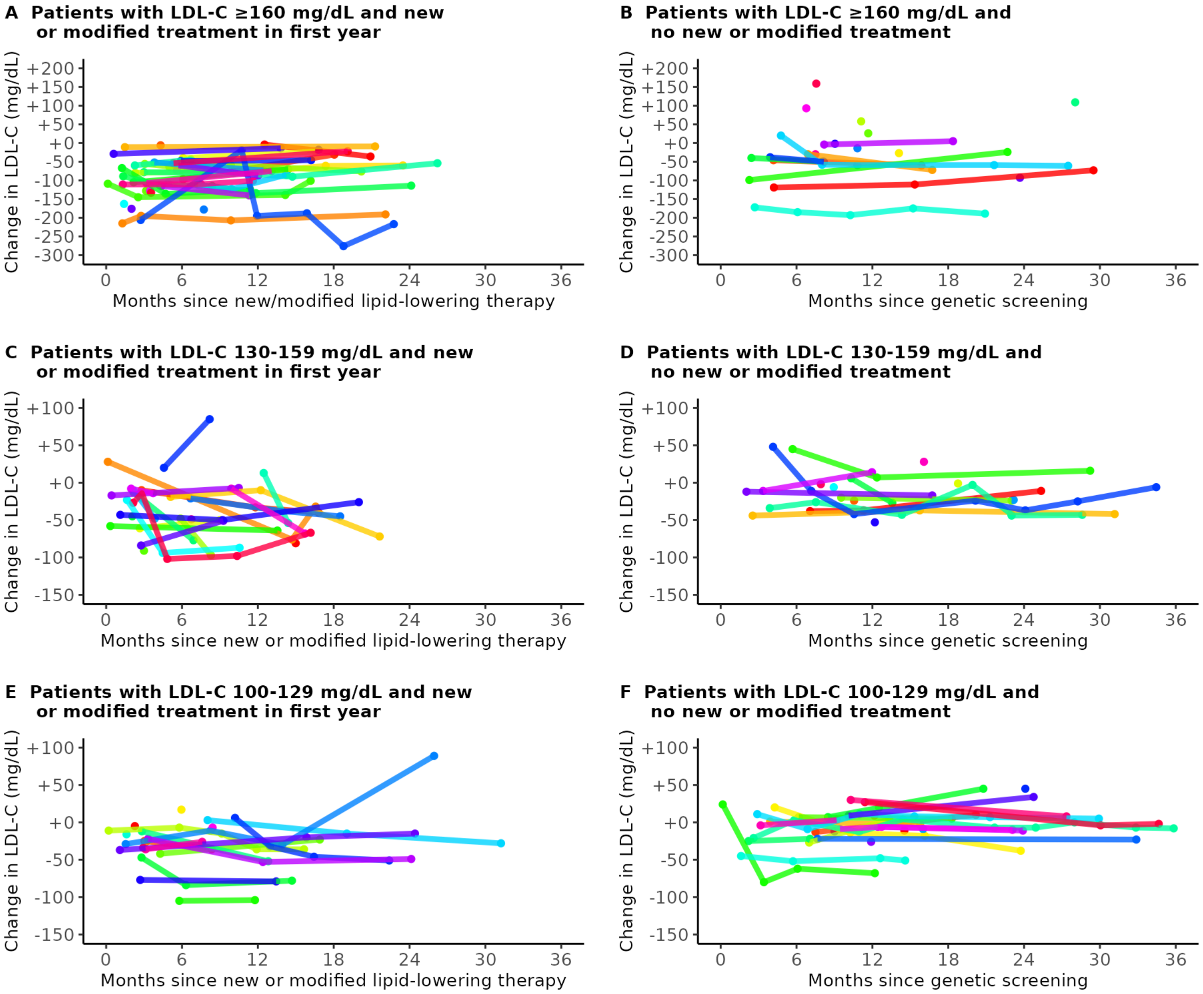


**Figure S3. Trajectories of LDL-C changes through up to 3 years after screening, stratified by baseline LDL-C concentration and lipid-lowering agent treatment status.** Includes patients with at least 1 LDL-C result spanning at least 6 months after genetic screening (for patients with no new/modified therapy) or after receipt of new/modified therapy (for those with new/modified therapy), in the first 3 years since screening. In those with no therapeutic addition/change (through up to 3 years), the y-axis is time since screening. In those with a therapeutic addition/change, the y-axis is time since first new/modified lipid-lowering therapy in the first year.

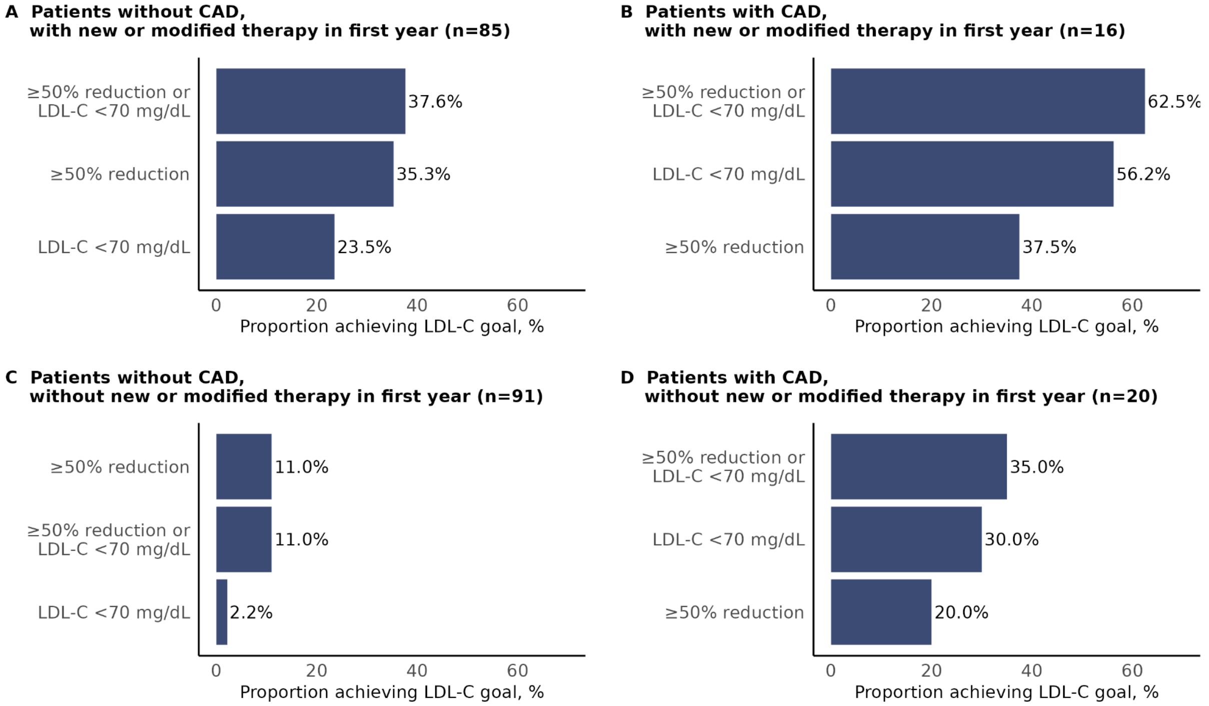


**Figure S4. Achievement of LDL-C target goals through up to 3 years post-genetic screening, stratified by CAD diagnosis status and treatment status.** Patients are included if they had a baseline LDL-C result available (through up to 1 year prior to genetic screening) and at least one follow-up LDL-C result. 18 patients with pre-screening LDL-C <70 mg/dL were excluded from this figure. One patient with the baseline LDL-C testing date within 1-60 days after screening was excluded because the new treatment occurred prior to the baseline LDL-C testing date. Among patients without a CAD diagnosis, p<0.001 for each of the three comparisons between patients with vs without new/modified therapy. Among patients with a CAD diagnosis, p=0.19, p=0.29, and p=0.21 for each of the three comparisons between patients with vs without new/modified therapy. CAD indicates coronary artery disease.
